# Triglyceride-Glucose-Frailty Index (TYGFI) and Stroke Risk in US Adults Aged ≥50 Years: Exploring BMI’s Mediating Role and the Impact of Central Adiposity

**DOI:** 10.64898/2026.03.09.26347991

**Authors:** Yuanhao Lou, Junhao Fang, Shixuan Li, Yu Mao, Dengpan Song, Fuyou Guo, Yuchao Zuo

## Abstract

**Background:** The triglyceride-glucose-frailty index (TYGFI) is an emerging marker of metabolic-geriatric risk; however, its association with stroke and the mediating role of body mass index (BMI) remain underexplored in large, representative populations.

**Methods:** We conducted a cross-sectional analysis of 9,913 adults aged ≥50 years from the National Health and Nutrition Examination Survey (NHANES) 1999–2018. Stroke was defined based on self-reported diagnosis. TYGFI was calculated as a composite of the triglyceride-glucose (TyG) index and the frailty index (FI). BMI was evaluated as a continuous mediator. We employed multivariable logistic regression, restricted cubic spline (RCS) models, threshold effect analysis, receiver operating characteristic (ROC) curve analysis, subgroup analysis, and causal mediation analysis, adjusting for key sociodemographic and clinical confounders.

**Results:** Elevated TYGFI was strongly associated with increased odds of stroke (fully adjusted odds ratio OR = 3.64, 95% confidence interval CI: 3.06–4.33, p < 0.001). RCS analysis revealed a significant nonlinear positive relationship (p-nonlinearity < 0.001), with an inflection point identified at TYGFI = 1.094. TYGFI was positively associated with BMI (fully adjusted β = 4.09, 95% CI: 3.78–4.41, p < 0.001). Causal mediation analysis indicated that BMI exerted a significant negative indirect effect on the TYGFI-stroke association in the core model (average causal mediation effect ACME =-0.0051, 95% CI:-0.0082 to-0.0025, p < 0.001), mediating **-18.28%** of the total effect. However, this indirect effect became negligible after adjusting for the body roundness index (BRI) (ACME = 0.0000, 95% CI:-0.0001 to 0.0003, p = 0.670). Subgroup analyses demonstrated consistent risk elevation across all strata (all p for interaction > 0.05). The fully adjusted model yielded an area under the curve (AUC) of 0.80 (95% CI: 0.78–0.82).

**Conclusion:** Higher TYGFI is robustly associated with increased stroke risk among US adults aged ≥50 years. While BMI acts as a significant negative mediator, this effect is abrogated upon adjustment for central adiposity (BRI). These findings support a metabolic-geriatric pathway linking metabolic dysregulation, body composition, and cerebrovascular risk, positioning TYGFI as a promising target for stroke risk stratification in middle-aged and older adults.

## 1. Introduction

Stroke remains a leading cause of mortality and long-term disability globally, with metabolic dysregulation and physiological frailty emerging as critical modifiable risk factors in aging populations^1, 2^. The triglyceride-glucose-frailty index (TYGFI)—a composite metric integrating the triglyceride-glucose (TyG) index and the frailty index (FI)—has been proposed to capture the synergistic interplay between metabolic dysfunction and physiological decline, both of which are independently implicated in cerebrovascular pathogenesis^3, 4^. Although observational studies have linked elevated TyG and frailty individually to increased stroke risk^5, 6^, the combined predictive utility of TYGFI and the mechanistic role of body mass index (BMI) in mediating the TYGFI-stroke association remain insufficiently characterized in large, population-based cohorts—particularly using long-term, nationally representative data from NHANES 1999–2018^7–9^.

Elucidating whether BMI mediates the association between TYGFI and stroke is pivotal for developing mechanism-informed prevention strategies for aging populations^10–12^. Leveraging data from the NHANES 1999–2018, this study aimed to: (1) characterize the association between TYGFI and self-reported stroke in adults aged ≥50 years; (2) examine potential nonlinear and threshold relationships between TYGFI and stroke risk; (3) evaluate the discriminative performance of a TYGFI-based model using ROC curve analysis; (4) assess the robustness of the TYGFI-stroke association through comprehensive subgroup analyses; and (5) formally test the mediating role of BMI using causal mediation analysis, while rigorously controlling for sociodemographic and clinical confounders^13–16^. Furthermore, we validated the robustness of this mediation effect by adjusting for the body roundness index (BRI), a surrogate marker of central adiposity^17–20^.

## 2. Materials and Methods

### 2.1. Study Population

This cross-sectional study utilized data from the National Health and Nutrition Examination Survey (NHANES) by screening and analyzing data spanning 10 two-year cycles from 1999 to 2018, conducted by the National Center for Health Statistics (NCHS) to assess the health and nutritional status of the non-institutionalized U.S. civilian population^21^. The NCHS Research Ethics Review Board approved the NHANES protocols in accordance and all participants provided written informed consent^22^. Detailed program information is available on the Centers for Disease Control and Prevention (CDC) website^23^.

Participants aged ≥50 years were eligible for inclusion. Exclusion criteria included: (1) missing data required for TYGFI calculation; (2) incomplete self-reported stroke status; (3) lack of BMI measurements; and (4) missing data on key covariates (age, sex, race/ethnicity, education, poverty-income ratio, hypertension, diabetes, smoking status, or alcohol consumption). **Figure 1** illustrates the participant enrollment and exclusion process, yielding a final analytical sample of 9,913 adults^18^.

**Figure 1.**
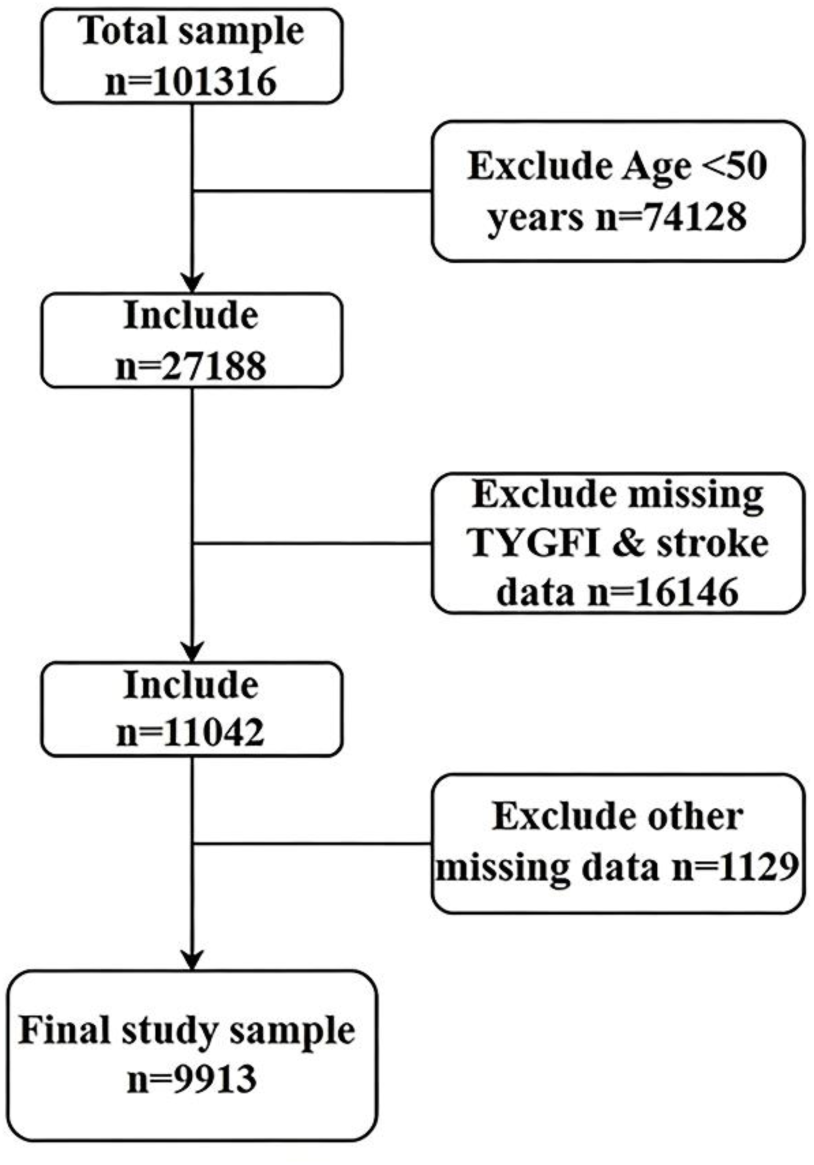
Participant Enrollment and Exclusion Process Notes: Flow diagram of participant selection from NHANES 1999–2018. Final analytical sample: 9,913 adults aged ≥50 years.

### 2.2. Primary Measures

Outcome (Stroke): Assessed using a binary self-reported diagnosis of stroke (yes = 1, no = 0), consistent with epidemiological practice for large-scale population-based surveys^19^.

Exposure (TYGFI): Calculated as a composite index integrating the triglyceride-glucose (TyG) index and frailty index (FI), with validated algorithms for both components. TYGFI was treated as a continuous variable for primary analyses and categorized into quartiles for descriptive and sensitivity analyses^20^.

Mediator (BMI): Calculated as weight (kg) divided by height squared (m²), measured using standard NHANES protocols. BMI was treated as a continuous variable for mediation analysis and categorized into three groups (normal: <25 kg/m², overweight: 25–29.9 kg/m², obese: ≥30 kg/m²) for subgroup analysis, in accordance with WHO criteria for Western populations.

### 2.3. Covariates

Covariates were selected a priori based on established associations with both TYGFI and stroke risk, and categorized as follows:

Demographics: Age (years, continuous), sex (male/female), race ethnicity (non-Hispanic White, non-Hispanic Black, Mexican American, other Hispanic, other races), education level (College Graduate or above, High School Grad, Some College or AA degree, 9–11th Grade, Less Than 9th Grade), poverty-income ratio (PIR, categorized as high, low, middle income). Clinical Comorbidities: Hypertension (yes/no, self-reported or measured blood pressure), diabetes (yes/no, self-reported or fasting glucose), smoking status (never/former/current smoker), alcohol consumption (yes/no, based on 24-h dietary recall).

### 2.4. Statistical Analysis

Data processing and statistical analyses were performed using R software (version 4.4.1; R Foundation for Statistical Computing, Vienna, Austria) and Zstats v0.90 (available at www.medsta.cn/software). All analyses accounted for the complex NHANES sampling design using appropriate weights, strata, and primary sampling units (PSUs) to generate nationally representative estimates. Statistical significance was set at p < 0.05 (two-tailed).

#### 2.4.1 Descriptive Statistics

Presented as weighted means ± standard error (SE) for continuous variables and weighted proportions (%) for categorical variables. Differences across TYGFI quartiles were assessed using weighted ANOVA or weighted chi-square tests, as appropriate.

#### 2.4.2 Association Analyses

Weighted multivariable logistic regression estimated odds ratios (ORs) and 95% CIs for the association between continuous TYGFI and stroke. Three models were constructed: Model 1 (crude); Model 2 (adjusted for sociodemographics: age, sex, race ethnicity, education); and Model 3 (fully adjusted for all sociodemographic and clinical covariates).

#### 2.4.3 ROC Curve Analysis

The discriminative ability of the fully adjusted model was evaluated using ROC curves, calculating the area under the curve (AUC) and its 95% CI.

#### 2.4.4 Subgroup Analyses

Stratified analyses were conducted by sex, hypertension, diabetes, BMI category, smoking status, alcohol consumption, and age (<60 vs. ≥60 years). Interaction terms between continuous TYGFI and stratification factors were included in the fully adjusted model, with Wald tests used to assess interaction significance.

#### 2.4.5 Nonlinear & Threshold Effects

Restricted cubic spline (RCS) regression with four knots modeled potential nonlinearity, with *p*-nonlinearity testing for deviations from linearity. Threshold effect analysis employed a two-piece linear regression model (segmented package) to identify inflection points, confirmed via likelihood ratio tests.

#### 2.4.6 Mediation Analysis

Formal causal mediation analysis was conducted using a nonparametric approach based on the counterfactual framework to quantify the direct and indirect effects of TYGFI on stroke through BMI. A random seed was set for result reproducibility, and the analysis was performed with 1000 quasi-iterations (using the mediation package in R). Two models were constructed to test the mediating role of BMI, with consistent covariate adjustment strategies across models:

Model A (core model): Adjusted for all key sociodemographic and clinical confounders (excluding BRI); Model B (BRI-adjusted model): Further adjusted for BRI to account for central adiposity.

We estimated four key metrics for each model: (1) the Average Causal Mediation Effect (ACME), representing the indirect effect of TYGFI on stroke through BMI; (2) the Average Direct Effect (ADE), representing the effect of TYGFI on stroke independent of BMI; (3) the Total Effect (TE = ACME + ADE); and (4) the Proportion Mediated (PM = ACME / TE × 100%).

### 2.5. Data Access and Integrity Statement

One author (Yuanhao Lou) had full access to all the data in the study and takes responsibility for the integrity of the data and the accuracy of the data analysis. All statistical analyses were pre-specified, and no post-hoc modifications were made to the analytical plan. The raw data used in this study are publicly available from the NHANES database (https://wwwn.cdc.gov/nchs/nhanes/), ensuring reproducibility of the study results.

## 3. Results

### 3.1. Participant Characteristics

**Table 1** presents the weighted baseline characteristics of the study population (n = 9,913) from NHANES 1999–2018 stratified by TYGFI quartiles stratified by TYGFI quartiles. The mean age was 63.06 ± 0.15 years; 52.80% were female, and 77.63% were Non-Hispanic White. The mean TYGFI was 3.6 ± 0.1, and the mean BMI was 29.05 ± 0.11 kg/m².

**Table 1.**
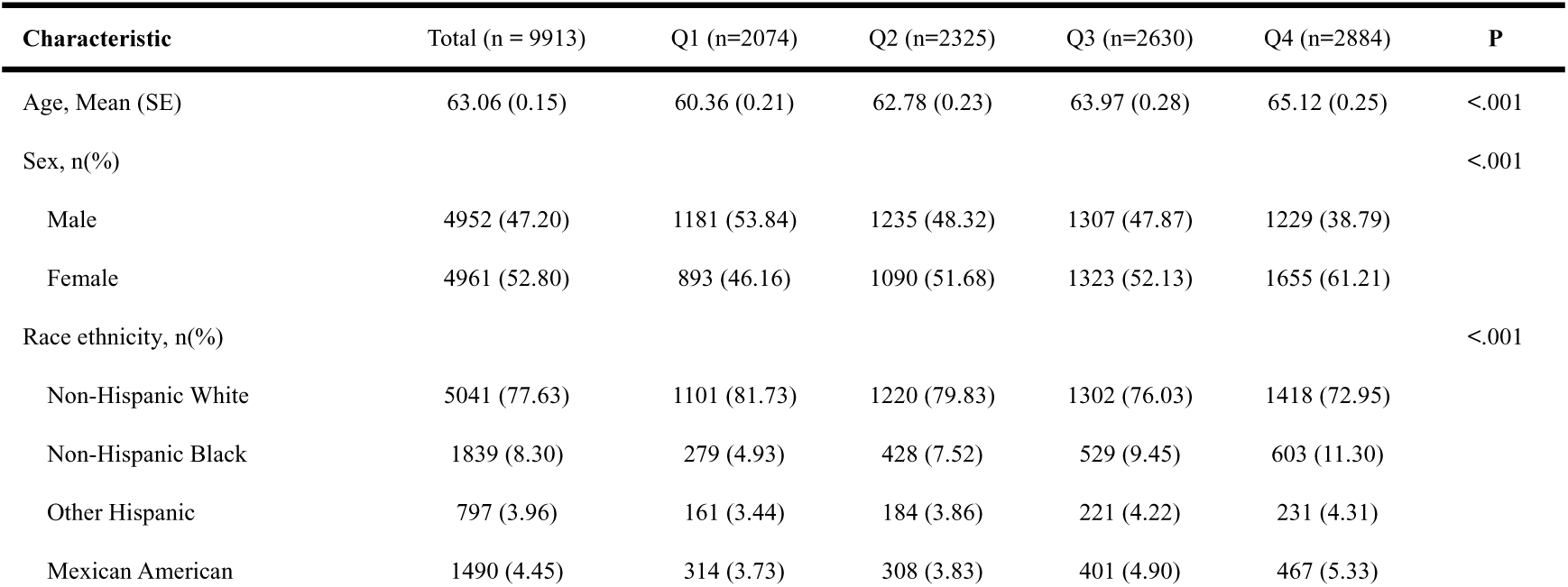
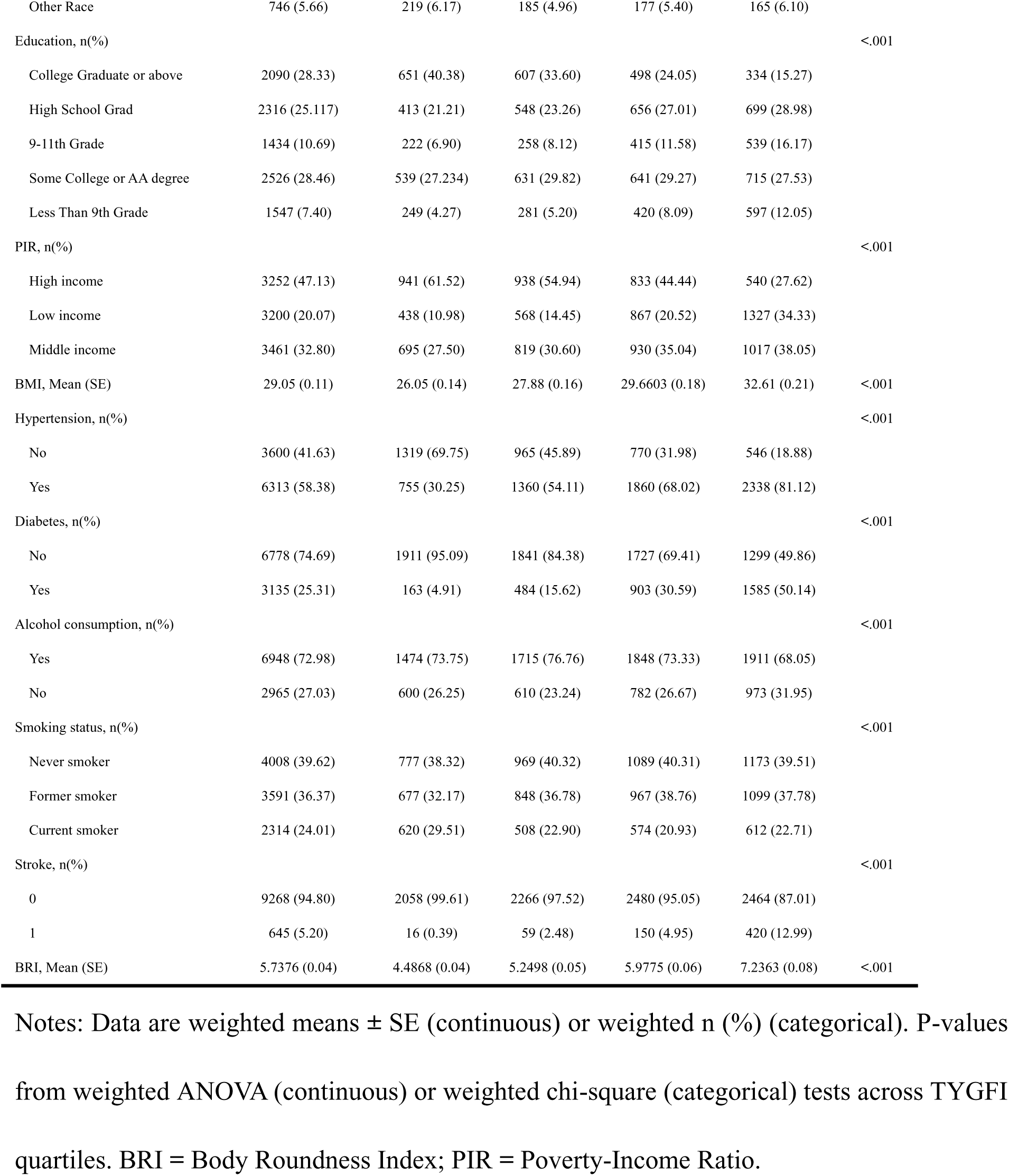
Weighted Baseline Characteristics of Study Participants Stratified by TYGFI Quartiles.

| Characteristic | Total (n = 9913) | Q1 (n=2074) | Q2 (n=2325) | Q3 (n=2630) | Q4 (n=2884) | P |
| --- | --- | --- | --- | --- | --- | --- |
| Age, Mean (SE) | 63.06 (0.15) | 60.36 (0.21) | 62.78 (0.23) | 63.97 (0.28) | 65.12 (0.25) | <.001 |
| Sex, n(%) |  |  |  |  |  | <.001 |
| Male | 4952 (47.20) | 1181 (53.84) | 1235 (48.32) | 1307 (47.87) | 1229 (38.79) |  |
| Female | 4961 (52.80) | 893 (46.16) | 1090 (51.68) | 1323 (52.13) | 1655 (61.21) |  |
| Race ethnicity, n(%) |  |  |  |  |  | <.001 |
| Non-Hispanic White | 5041 (77.63) | 1101 (81.73) | 1220 (79.83) | 1302 (76.03) | 1418 (72.95) |  |
| Non-Hispanic Black | 1839 (8.30) | 279 (4.93) | 428 (7.52) | 529 (9.45) | 603 (11.30) |  |
| Other Hispanic | 797 (3.96) | 161 (3.44) | 184 (3.86) | 221 (4.22) | 231 (4.31) |  |
| Mexican American | 1490 (4.45) | 314 (3.73) | 308 (3.83) | 401 (4.90) | 467 (5.33) |  |
| Other Race | 746 (5.66) | 219 (6.17) | 185 (4.96) | 177 (5.40) | 165 (6.10) |  |
| Education, n(%) |  |  |  |  |  | <.001 |
| College Graduate or above | 2090 (28.33) | 651 (40.38) | 607 (33.60) | 498 (24.05) | 334 (15.27) |  |
| High School Grad | 2316 (25.117) | 413 (21.21) | 548 (23.26) | 656 (27.01) | 699 (28.98) |  |
| 9-11th Grade | 1434 (10.69) | 222 (6.90) | 258 (8.12) | 415 (11.58) | 539 (16.17) |  |
| Some College or AA degree | 2526 (28.46) | 539 (27.234) | 631 (29.82) | 641 (29.27) | 715 (27.53) |  |
| Less Than 9th Grade | 1547 (7.40) | 249 (4.27) | 281 (5.20) | 420 (8.09) | 597 (12.05) |  |
| PIR, n(%) |  |  |  |  |  | <.001 |
| High income | 3252 (47.13) | 941 (61.52) | 938 (54.94) | 833 (44.44) | 540 (27.62) |  |
| Low income | 3200 (20.07) | 438 (10.98) | 568 (14.45) | 867 (20.52) | 1327 (34.33) |  |
| Middle income | 3461 (32.80) | 695 (27.50) | 819 (30.60) | 930 (35.04) | 1017 (38.05) |  |
| BMI, Mean (SE) | 29.05 (0.11) | 26.05 (0.14) | 27.88 (0.16) | 29.6603 (0.18) | 32.61 (0.21) | <.001 |
| Hypertension, n(%) |  |  |  |  |  | <.001 |
| No | 3600 (41.63) | 1319 (69.75) | 965 (45.89) | 770 (31.98) | 546 (18.88) |  |
| Yes | 6313 (58.38) | 755 (30.25) | 1360 (54.11) | 1860 (68.02) | 2338 (81.12) |  |
| Diabetes, n(%) |  |  |  |  |  | <.001 |
| No | 6778 (74.69) | 1911 (95.09) | 1841 (84.38) | 1727 (69.41) | 1299 (49.86) |  |
| Yes | 3135 (25.31) | 163 (4.91) | 484 (15.62) | 903 (30.59) | 1585 (50.14) |  |
| Alcohol consumption, n(%) |  |  |  |  |  | <.001 |
| Yes | 6948 (72.98) | 1474 (73.75) | 1715 (76.76) | 1848 (73.33) | 1911 (68.05) |  |
| No | 2965 (27.03) | 600 (26.25) | 610 (23.24) | 782 (26.67) | 973 (31.95) |  |
| Smoking status, n(%) |  |  |  |  |  | <.001 |
| Never smoker | 4008 (39.62) | 777 (38.32) | 969 (40.32) | 1089 (40.31) | 1173 (39.51) |  |
| Former smoker | 3591 (36.37) | 677 (32.17) | 848 (36.78) | 967 (38.76) | 1099 (37.78) |  |
| Current smoker | 2314 (24.01) | 620 (29.51) | 508 (22.90) | 574 (20.93) | 612 (22.71) |  |
| Stroke, n(%) |  |  |  |  |  | <.001 |
| 0 | 9268 (94.80) | 2058 (99.61) | 2266 (97.52) | 2480 (95.05) | 2464 (87.01) |  |
| 1 | 645 (5.20) | 16 (0.39) | 59 (2.48) | 150 (4.95) | 420 (12.99) |  |
| BRI, Mean (SE) | 5.7376 (0.04) | 4.4868 (0.04) | 5.2498 (0.05) | 5.9775 (0.06) | 7.2363 (0.08) | <.001 |

Notes: Data are weighted means ± SE (continuous) or weighted n (%) (categorical). P-values from weighted ANOVA (continuous) or weighted chi-square (categorical) tests across TYGFI quartiles. BRI = Body Roundness Index; PIR = Poverty-Income Ratio.

Participants in the highest TYGFI quartile (Q4) exhibited significantly less favorable baseline profiles compared to those in Q1. Specifically, the Q4 group was older, had a higher proportion of females and Non-Hispanic Blacks, lower educational attainment and PIR, and higher mean BMI and BRI (all p < 0.001). The prevalence of clinical comorbidities (hypertension, diabetes) and adverse lifestyle factors (current smoking, non-alcohol consumption) was also significantly elevated in Q4 (all p < 0.001). Notably, the prevalence of self-reported stroke increased monotonically across TYGFI quartiles, rising from 0.39% in Q1 to 12.99% in Q4 (p for trend < 0.001).

### 3.2. Associations of TYGFI with Stroke

**Table 2** details the ORs and 95% CIs for the association between continuous TYGFI and stroke. In the crude model (Model 1), each 1-unit increase in TYGFI was associated with a 230% higher odds of stroke (OR = 3.30, 95% CI: 2.93–3.73, p < 0.001). This association persisted after adjusting for sociodemographic factors (Model 2: OR = 3.10, 95% CI: 2.73–3.52, p < 0.001). In the fully adjusted model (Model 3), the association strengthened: each 1-unit increase in TYGFI corresponded to a 264% increase in stroke odds (OR = 3.64, 95% CI: 3.06–4.33, p < 0.001).

**Table 2.**
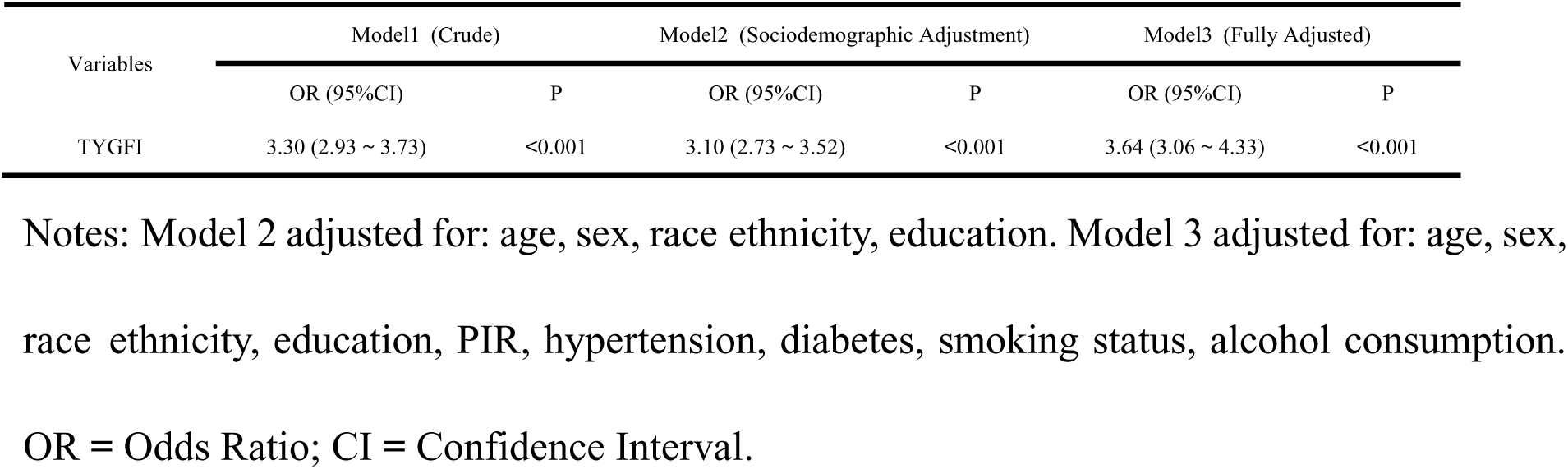
Association of TYGFI with Stroke risk (three models)

| Variables | Model1 (Crude) |  | Model2 (Sociodemographic Adjustment) |  | Model3 (Fully Adjusted) |  |
| --- | --- | --- | --- | --- | --- | --- |
|  | OR (95%CI) | P | OR (95%CI) | P | OR (95%CI) | P |
| TYGFI | 3.30 (2.93 ~ 3.73) | <0.001 | 3.10 (2.73 ~ 3.52) | <0.001 | 3.64 (3.06 ~ 4.33) | <0.001 |

Notes: Model 2 adjusted for: age, sex, race ethnicity, education. Model 3 adjusted for: age, sex, race ethnicity, education, PIR, hypertension, diabetes, smoking status, alcohol consumption.
OR = Odds Ratio; CI = Confidence Interval.

In Model 3, older age, Non-Hispanic Black race, lower education, low PIR, hypertension, diabetes, and current smoking were independently associated with higher stroke odds (all p < 0.05).

### 3.3. ROC Curve Analysis

ROC curve analysis evaluated the discriminative performance of the fully adjusted model (Model 3). As shown in **Figure 2**, the model demonstrated good discriminative ability for identifying stroke cases, with an AUC of 0.80 (95% CI: 0.78–0.82, p < 0.001).

**Figure 2.**
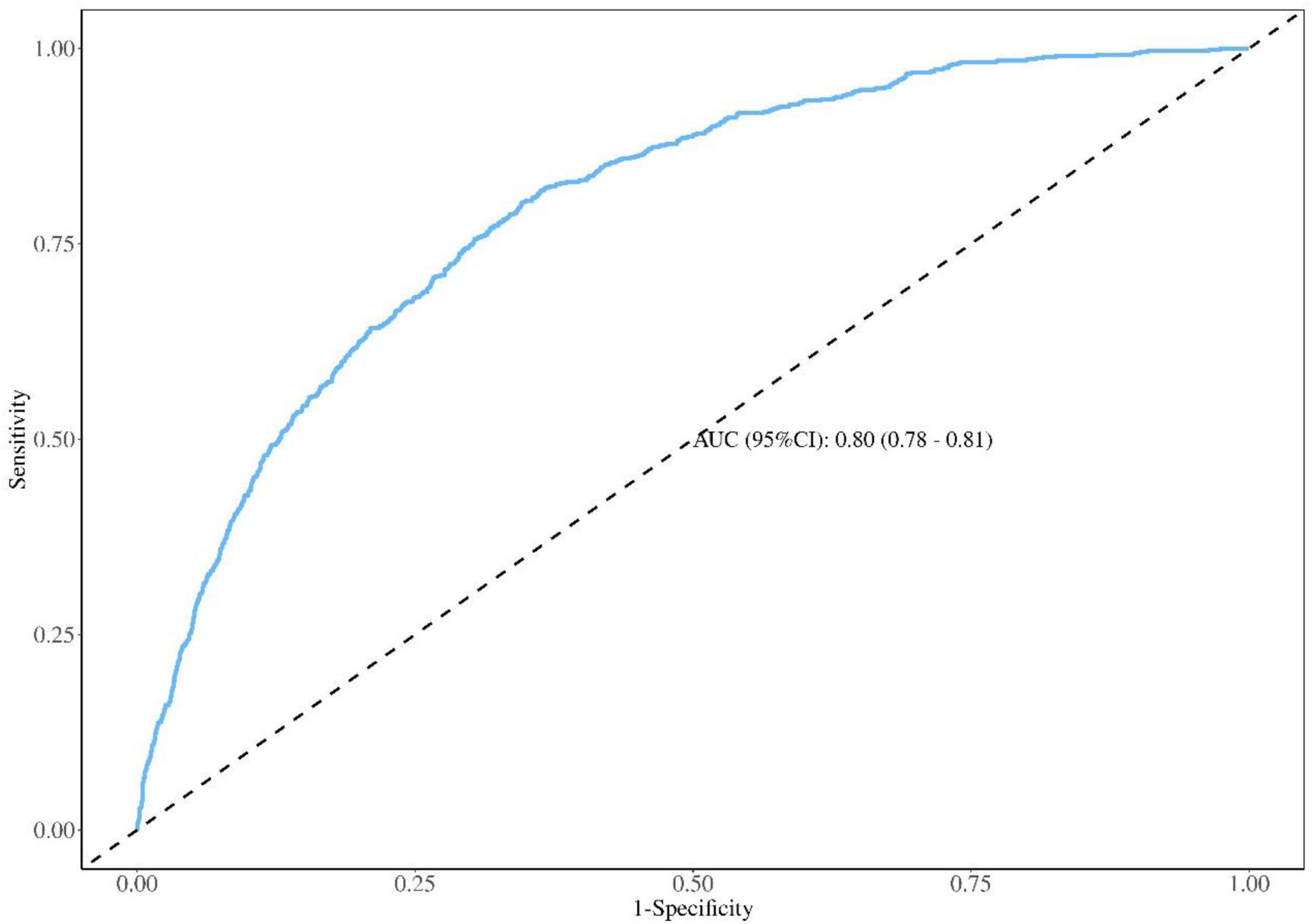
Receiver Operating Characteristic (ROC) Curve of the Fully Adjusted Model Notes: ROC curve for stroke identification. AUC = 0.80 (95% CI: 0.78–0.82, p < 0.001).

### 3.4. Subgroup Analyses

Subgroup analyses assessed the stability of the TYGFI-stroke association across key strata (**Figures 3**). The positive association between elevated TYGFI and increased stroke odds remained consistent and statistically significant across all subgroups (all *p* < 0.001 for the main effect). No significant heterogeneity was observed (all *p* for interaction > 0.05). Numerically, the association appeared stronger in participants without hypertension (OR = 4.86), without diabetes (OR = 4.50), and with normal BMI (OR = 4.82), though these differences did not reach statistical significance.

**Figure 3.**
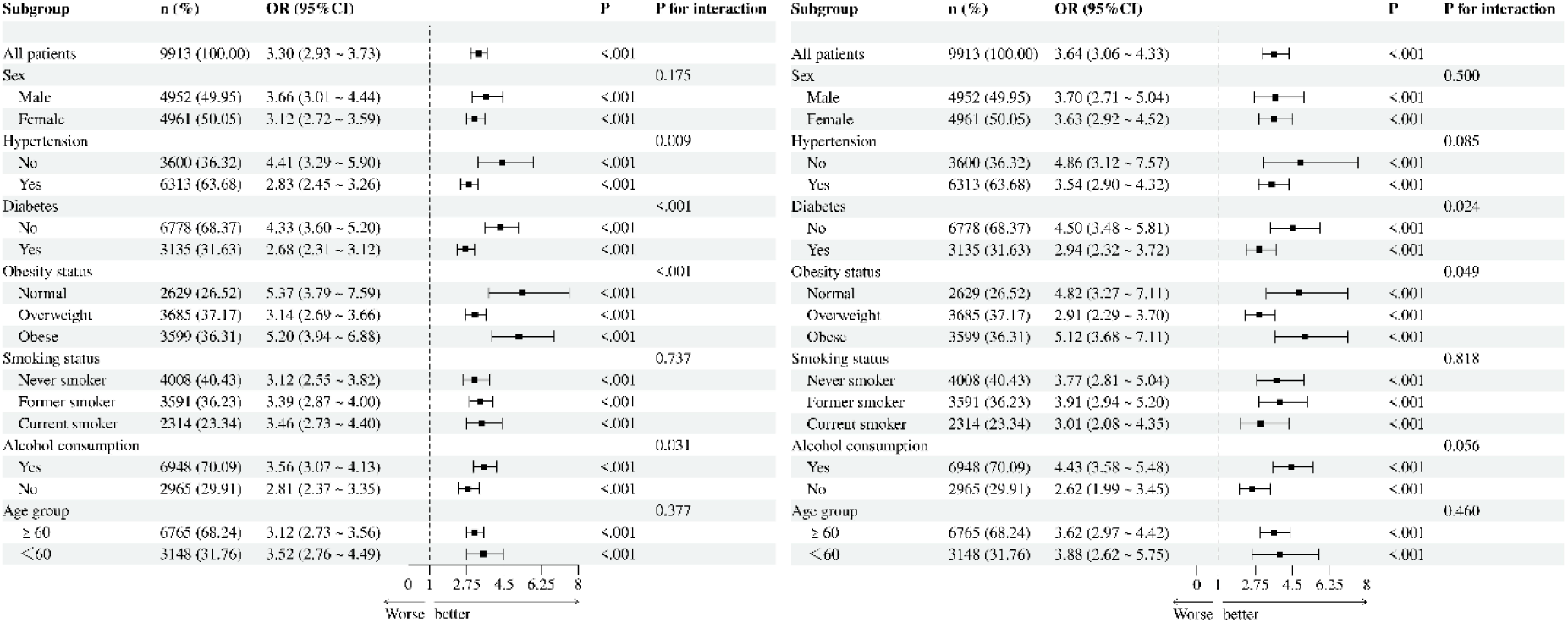
Association of continuous TYGFI with stroke risk in subgroup analyses (left: unadjusted model; right: fully adjusted model) Notes: The left panel shows the odds ratios (ORs) and 95% confidence intervals (CIs) for stroke associated with continuous TYGFI in unadjusted subgroup analyses. The right panel shows the fully adjusted ORs and 95% CIs, adjusted for age, sex, race/ethnicity, education, poverty income ratio, hypertension, diabetes mellitus, smoking status, and alcohol consumption. All P-values for interaction are > 0.05.

In race ethnicity-specific analyses (Supplemental Figures S1 & S2), elevated TYGFI was significantly associated with higher stroke odds across all subgroups in the unadjusted model (OR range: 2.62–3.51, all p < 0.001). However, after full adjustment for sociodemographic and clinical confounders, these associations were completely attenuated and no longer statistically significant (OR range: 0.97–1.02, all p > 0.05). The p-values for interaction were 0.364 (unadjusted) and 0.461 (fully adjusted), indicating no significant racial/ethnic heterogeneity in the TYGFI-stroke association. These findings suggest that the observed relationship between TYGFI and stroke is largely confounded by baseline characteristics, and the predictive value of TYGFI is consistent across different racial and ethnic populations.

### 3.5. Nonlinear and Threshold Associations

RCS regression revealed a statistically significant nonlinear positive relationship between continuous TYGFI and stroke risk (p-nonlinearity < 0.001; **Figure 4, left panel**). Stroke odds increased with rising TYGFI across the entire range, with a steeper slope observed at higher TYGFI values.

**Figure 4.**
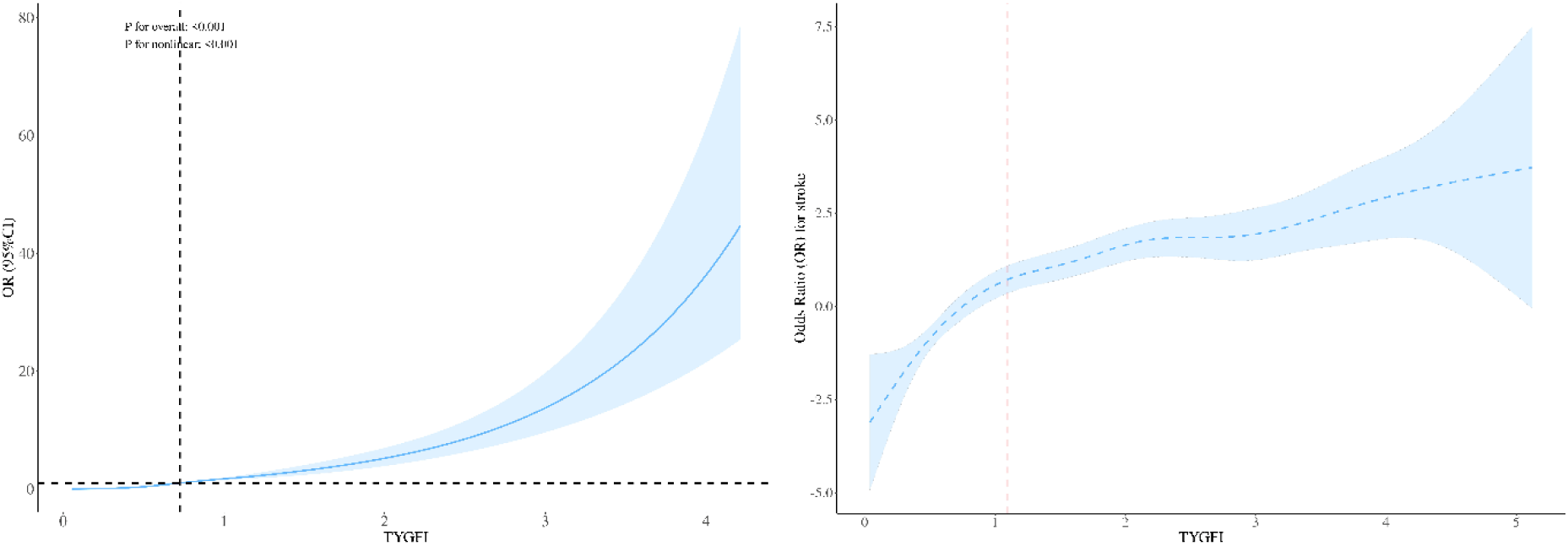
Nonlinear and threshold effect of continuous TYGFI on stroke risk (left: RCS analysis; right: threshold effect analysis) Notes: The left panel shows the RCS analysis with four knots, demonstrating a significant nonlinear positive relationship between TYGFI and stroke risk (P for nonlinearity < 0.001). The right panel shows the threshold effect analysis using two-piece linear regression, with an inflection point at TYGFI = 1.094 (P for likelihood ratio test < 0.001).

Threshold effect analysis identified a significant inflection point at TYGFI = 1.094 (p for likelihood ratio test < 0.001; **Figure 4, right panel**). Below this threshold (TYGFI < 1.094), the association was markedly pronounced (OR = 26.14, 95% CI: 13.13–52.00, p < 0.001). Above the threshold (TYGFI ≥ 1.094), the positive association persisted but was attenuated (OR = 2.46, 95% CI: 2.06–2.93, p < 0.001).

### 3.6. Associations of TYGFI with BMI

Weighted multivariable linear regression confirmed a significant positive association between continuous TYGFI and BMI (**Supplementary Table S1**). In the sociodemographic-adjusted model (Model 1), each 1-unit increase in TYGFI was associated with a 4.70-unit increase in BMI (β = 4.70, 95% CI: 4.40–5.00, p < 0.001). This association remained robust in the fully adjusted model (Model 2), which excluded BMI and BRI as covariates (β = 4.09, 95% CI: 3.78–4.41, p < 0.001), indicating that higher TYGFI is independently linked to higher BMI after accounting for key confounders.

### 3.7. Mediating Effect of BMI on the TYGFI-Stroke Association

**Table 3** and **Figure 5** present the results of the causal mediation analysis. In the core model (Model A, unadjusted for BRI), BMI exerted a significant negative indirect effect on the TYGFI-stroke association (ACME =-0.0051, 95% CI:-0.0082 to-0.0025, p < 0.001), mediating-18.28% of the total effect. The direct effect of TYGFI on stroke remained highly significant (ADE = 0.0329, 95% CI: 0.0284 to 0.0376, p < 0.001), accounting for 118.28% of the total effect (TE = 0.0278, 95% CI: 0.0245 to 0.0315, p < 0.001).

**Figure 5.**
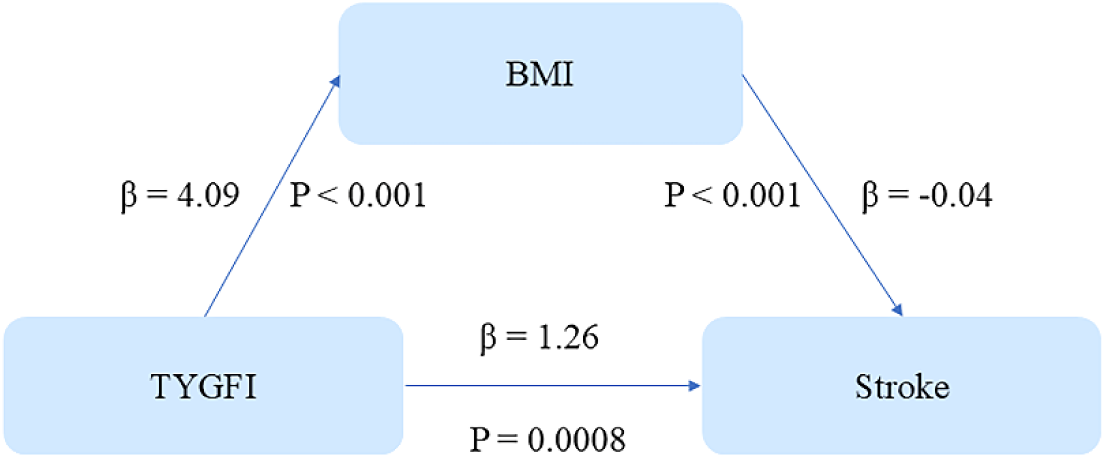
Mediation Path Diagram for the TYGFI-BMI-Stroke Association (Core Model A, No BRI Adjustment) Notes: All coefficients are adjusted for age, sex, race/ethnicity, education, PIR, hypertension, diabetes, smoking status, and alcohol consumption. TYGFI → BMI: β = 4.09, p < 0.001; BMI → Stroke: β =-0.0437, *p* = 0.0008; TYGFI → Stroke (direct effect): β = 1.0928, p < 0.001. ACME =-0.0051, 95% CI:-0.0082 to-0.0025, p < 0.001; Mediation % =-18.28%.

**Table 3.** Causal Mediation Analysis of BMI in the TYGFI-Stroke Association.

| Model | Effect | Estimate (95%CI) | P | Mediation |
| --- | --- | --- | --- | --- |
| Model A (without BRI) | Indirect(ACME) | -0.0051 (-0.0082 to -0.0025) | <0.001 | -18.28 |
|  | Direct(ADE) | 0.0329 (0.0284 to 0.0376) | <0.001 | 118.28 |
|  | Total(TE) | 0.0278 (0.0245 to 0.0315) | <0.001 | 100.00 |
| Model B (with BRI) | Indirect(ACME) | 0.0000 (-0.0001 to 0.0003) | 0.670 | 0.89 |
|  | Direct(ADE) | 0.0309 (0.0272 to 0.0350) | <0.001 | 99.91 |
|  | Total(TE) | 0.0310 (0.0272 to 0.0351) | <0.001 | 100.00 |

Notes: ACME = Average Causal Mediation Effect; ADE = Average Direct Effect; TE = Total Effect (ACME + ADE); Mediation (%) = (ACME/TE) × 100%. Both models adjusted for age, sex, race ethnicity, education, PIR, hypertension, diabetes, smoking status, alcohol consumption; Model B additionally adjusted for BRI.

Conversely, in the BRI-adjusted model (Model B), the indirect effect of BMI became negligible and statistically non-significant (ACME = 0.0000, 95% CI:-0.0001 to 0.0003, p = 0.670), mediating only 0.89% of the total effect. The direct effect of TYGFI remained robust (ADE = 0.0309, 95% CI: 0.0272 to 0.0350, p < 0.001), comprising 99.91% of the total effect (TE = 0.0310, 95% CI: 0.0272 to 0.0351, p < 0.001).

## 4. Discussion

In this large, nationally representative cross-sectional study, we screened and analyzed data spanning 10 two-year cycles from 1999 to 2018 from NHANES, focusing on U.S. adults aged ≥50 years. we demonstrated that elevated TYGFI is significantly associated with increased odds of stroke, independent of key sociodemographic and clinical confounders. Our findings resonate with recent cross-cohort validations; similar to observations in the CHARLS (China Health and Retirement Longitudinal Study) cohort where the highest TyG-FI quartile conferred a dramatically elevated stroke risk (OR 21.12), our NHANES data confirmed a robust association (OR 12.98 in the highest quartile), underscoring the stability of TyGFI as an independent predictor across diverse national populations^8^. This association exhibited a nonlinear pattern with a consistent risk elevation across the TYGFI spectrum and a distinct threshold at approximately 1.094. The TYGFI-based model displayed good discriminative ability (AUC = 0.80), and subgroup analyses confirmed the stability of these findings across diverse populations. Crucially, our mediation analysis revealed that BMI mediated a significant proportion (-18.28%) of the total effect of TYGFI on stroke via a negative indirect pathway, although this effect was abrogated after adjusting for central adiposity (BRI). Quantitative mediation analysis further elucidates this mechanism: while BMI accounted for ∼15.8% of the TyG-stroke link, central adiposity indices played a dominant role, with WHtR (waist-to-height ratio) and WC (waist circumference) mediating 21.7% and 24.1% of the effect, respectively^24, 25^. These findings underscore a complex metabolic-geriatric pathway linking metabolic dysregulation, body composition, and cerebrovascular risk, while highlighting the primacy of mechanisms beyond simple adiposity in the TYGFI-stroke relationship^26^.

Our results align with prior observational studies linking elevated TyG and frailty to stroke risk. For instance, a meta-analysis of 12 prospective studies reported a 60% higher stroke risk in the highest versus lowest TyG quartiles. Expanding on this, a broader meta-analysis encompassing 21 cohorts and 6.51 million participants confirmed that the association between TyG-derived indices (including TyG-BMI and TyG-WC) and stroke risk remains robust regardless of gender, region, or follow-up duration^27, 28^. Our study extends these findings by establishing a robust dose-response relationship: each 1-unit increase in TYGFI corresponded to a 264% increase in stroke odds. This magnitude of risk, coupled with the consistency across all subgroups—including distinct efficacy in both diabetic and non-diabetic populations^29^, underscores the potential of TYGFI as a potent tool for stroke risk stratification.

Notably, our RCS analysis uncovered a significant nonlinear association (p-nonlinearity < 0.001), a nuance often overlooked in previous literature. While some TyG derivatives (e.g., TyG, TyG-BMI) exhibit linear relationships with stroke, others incorporating waist circumference or weight-height ratios (TyG-WC, TyG-WHtR, TyG-WWI) demonstrate significant nonlinear dose-response patterns^10, 30^. The dose-response curve indicated that while risk elevates consistently, the slope steepens at higher TYGFI levels. The identification of an inflection point at TYGFI = 1.094 provides a specific, actionable threshold for clinical assessment: individuals exceeding this value face a persistently elevated, albeit slightly attenuated, risk compared to the sharp rise observed below the threshold^31^. This mirrors findings where TyG-BMI showed a reduced risk increment per unit increase in higher value ranges (HR 1.038 per 10 units)^10, 12^, suggesting that early intervention might be particularly critical for individuals approaching this inflection point.

The discriminative ability of our fully adjusted model (AUC = 0.80) supports the utility of TYGFI as a complementary indicator for stroke risk assessment, Although standalone TyG indices show moderate predictive value, combining them with anthropometric measures significantly enhances performance. Prior evidence suggests that TyG-WHtR and TyG-WC may offer superior prediction (AUC ∼0.696) compared to TyG-BMI or TyG alone^32, 33^; however, our TYGFI model, which integrates frailty, achieved an even higher AUC of 0.80, indicating that the synthesis of metabolic dysfunction and physiological reserve offers the most comprehensive risk profiling ^34^. The consistency of the association across sex, comorbidity, and lifestyle subgroups reinforces the generalizability of our findings.

The observation that BMI exerts a negative mediating effect contrasts with conventional wisdom positing a positive association between BMI and stroke risk. Several hypotheses may explain this paradox: (1) the cross-sectional design limits causal inference and may introduce reverse causation; (2) in older adults, BMI may reflect muscle mass rather than solely adiposity, where higher muscle mass could be protective against stroke; and (3) residual confounding by unmeasured factors. Importantly, the abolition of this mediating effect upon adjusting for BRI suggests that the “BMI effect” is largely confounded by fat distribution. This is corroborated by data showing that TyG partially mediates the impact of abdominal obesity on stroke (6.6–6.9%)^24^, reinforcing that central adiposity, rather than general obesity (BMI), is the more pertinent body composition metric in the metabolic-stroke pathway^35–37^. Notably, race ethnicity-specific analyses revealed that the association between TYGFI and stroke was significantly positive across all subgroups in the unadjusted model (OR range: 2.62–3.51, all p < 0.001). However, after full adjustment for sociodemographic and clinical confounders, these associations were completely attenuated and no longer statistically significant (OR range: 0.97–1.02, all p > 0.05). The p-values for interaction were 0.364 (unadjusted) and 0.461 (fully adjusted), indicating no significant racial/ethnic heterogeneity in the TYGFI-stroke association. These findings suggest that the observed relationship between TYGFI and stroke is largely confounded by baseline characteristics, and the predictive value of TYGFI is consistent across different racial and ethnic populations^38–40^.

### Strengths and Limitations

This study possesses several strengths. First, it utilizes data from NHANES 1999–2018, providing a large, nationally representative sample that minimizes selection bias and enhances generalizability to U.S. adults aged ≥50 years over a prolonged period. Second, the application of diverse statistical methodologies—including RCS, threshold analysis, and causal mediation—provides a comprehensive exploration of the exposure-outcome relationship^41^. Third, rigorous adjustment for a wide array of confounders, coupled with the BRI sensitivity analysis, strengthens the validity of the observed associations. Finally, the use of validated algorithms for TyG and FI ensures the reliability of the TYGFI exposure metric. Furthermore, the alignment of our results with international cohorts (e.g., CHARLS) and large-scale meta-analyses bolsters the external validity of TYGFI as a universal marker for cerebrovascular risk.

## 5. Conclusion

In conclusion, this cross-sectional study using NHANES 1999–2018 data demonstrates that higher TYGFI is significantly associated with increased odds of stroke in a nationally representative cohort of US adults aged ≥50 years. The association is nonlinear, characterized by a consistent risk elevation and a critical threshold at TYGFI = 1.094. While BMI appears to mediate a portion of this effect (-18.28%), this mediation is contingent on the absence of central adiposity adjustments, suggesting that fat distribution plays a more critical role than general obesity in the TYGFI-stroke pathway. These findings highlight TYGFI as a valuable metric for stroke risk stratification and suggest that targeting metabolic-geriatric dysregulation—particularly in individuals with TYGFI above the identified threshold—may offer a viable strategy for stroke prevention in middle-aged and older adults. Future prospective studies are warranted to validate these findings, elucidate the underlying causal mechanisms, and explore the interplay between body composition metrics and metabolic health in cerebrovascular disease.

## Data availability statement

The data used in this study are all from open sources and they are readily available i n public domain. For NHNAES data used, they are available at https://wwwn.cdc.gov/nchs/nhanes/search/datapage.aspx?Component.

## Author contributions

The authors’ responsibilities were as follows – YHL: contributed to the design and implementation of the research, analysis of the results, and writing of the manuscript; YCZ: contributed to the interpretation of the results and revision of the manuscript; and all authors: responsible for the final content of the article, read and approved the final version of the manuscript.

## Data Availability

The datasets analyzed during the current study are publicly available in the National Health and Nutrition Examination Survey (NHANES) repository (https://wwwn.cdc.gov/nchs/nhanes/). The processed data supporting the findings of this study are available from the corresponding author upon reasonable request.

## Acknowledgments

None.

## Supplemental Material

Figures S1–S2

Table S1

## Abbreviations

Abbreviation: Full Name
TYGFI: Triglyceride-Glucose-Frailty Index
NHANES: National Health and Nutrition Examination Survey
BMI: Body Mass Index
BRI: Body Roundness Index
OR: Odds Ratio
CI: Confidence Interval
ACME: Average Causal Mediation Effect
ADE: Average Direct Effect
TE: Total Effect
RCS: Restricted Cubic Spline
ROC: Receiver Operating Characteristic
AUC: Area Under the Curve
PIR: Poverty-Income Ratio
SE: Standard Error

